# An Investigation into Discomfort and Fatigue Related to the Wearing of an EEG Neurofeedback Headset

**DOI:** 10.1101/2023.02.16.23284115

**Authors:** Simon Hanzal, Lucie Tvrda, Monika Harvey

## Abstract

Self-regulating brain activity using electroencephalography (EEG) neurofeedback has the potential to improve cognitive functions, rehabilitate motor control and reduce chronic fatigue. Nonetheless, user experience and factors which may interfere with the beneficial effects of neurofeedback are still under researched. This preliminary study aimed to investigate whether wearing an EEG neurofeedback recommended headset for 1 hour induced significant physical discomfort and fatigue. Data were obtained from a standard visual analogue scale questionnaire and a newly developed EEG headset discomfort (EEGhd) questionnaire. 21 participants (12 in the experimental (headset) and 9 in the control (electrodes only) group) watched a nature documentary video while their brain signals were recorded. They completed the set of questionnaires before and after the video, while wearing the EEG headset (or electrodes). A two-sample t-test revealed that the experimental group experienced significantly higher EEGhd than the controls (p < .001). Participants reported the onset of discomfort after approximately 25 minutes. These results highlight the importance of assessing user experience and accounting for physical discomfort when designing an EEG neurofeedback study.

## Introduction

Electroencephalographic (EEG) neurofeedback is a non-invasive rehabilitation tool used to treat long-term problems with cognitive function. It utilises visual training to assist with self-regulating brain activity at specific frequencies. It has been used for the rehabilitation of motor control after stroke (Foong et al., 2020), as well as for the enhancement of cognitive functions and to reduce chronic fatigue (Lucktar-Flude & Groll, 2015).

Although some research highlights the beneficial effects of neurofeedback, further evidence is needed to determine the useability of EEG neurofeedback headsets. Marzbani et al. (2016) suggested the use of a bipolar montage of active electrodes, where the signal is recorded by 2 electrodes per channel (active and reference). Ear clips were suggested for the placement of the reference electrodes. Other researchers have pointed out the importance of securing electrodes with enough pressure by using headbands that ensure good contact with the participant’s scalp and thus sufficient quality of the EEG signal to guide online neurofeedback (Verwulgen et al., 2018; Hairston et al., 2014). Headbands reduce external artifacts and are intended to remain comfortable for the participant. Jochumsen et al. (2020) found headbands to be the least complex, fastest to mount and most preferred by patients and therapists alike.

On the other hand, EEG headbands have been associated with some level of pressure (Verwulgen et al., 2018). Participants with larger head sizes often report higher discomfort due to increased pressure of the headset (Nijboer et al., 2015). Moreover, consistent pressure may also increase this discomfort over time (Hairston et al., 2014). Pei et al. (2018) emphasised the importance of reducing this detected wearing discomfort when using EEG neurofeedback devices. However, most previous EEG useability studies have simply focused on the functionality of the device and its signal quality (Ries et al., 2014), despite the fact that some studies have found that wearing comfort and visual appearance of the electrodes has an impact on participants’ overall experience, and their ability to focus on the task (Nikulin et al., 2010; Raduntz & Meffert, 2019). Raduntz & Meffert (2019) compared participants’ experience across 7 EEG devices. They found that these EEG devices differed significantly in wearing comfort, with pin electrodes and rigid headsets being the most uncomfortable. Moreover, wearing comfort was a key factor in the participants’ willingness to use the device.

EEG studies have also found that physical pain and discomfort are associated with changes in the power of the different frequency bands, such as increases in Beta power (De Pascalis et al., 2019) and decreases in Alpha power (Shao et al., 2012). Physical discomfort and fatigue interfere also with cognitive task performance (DenHartog & Koerhuis, 2017; Myrden & Chau, 2015). Therefore, it is possible that the physical discomfort associated with wearing an EEG headset with a headband during a neurofeedback rehabilitation session, may be masking patient’s true means of changing EEG power, decreasing their ability to focus on a task and thus an interference with the potential treatment effects.

The aim of this single-blinded preliminary study was to investigate aspects of the EEG user experience that could potentially interfere with the effect of future neurofeedback rehabilitation of post-stroke fatigue. Specifically, the study examined whether the recommended neurofeedback EEG headset including a headband and ear clips, caused greater physical discomfort and fatigue than a minimal EEG headset with electrodes alone. Moreover, to date, there is no standard measure for assessing discomfort related to EEG headsets. Therefore, this study hypothesised that participants wearing the EEG neurofeedback headset including a headband and ear clips would experience 1) higher change in an EEG headset discomfort (EEGhd) score obtained from a new EEGhd measurement tool and 2) a higher state fatigue change than control participants wearing EEG electrodes alone without a headset. Furthermore, it was expected that a larger head diameter would be associated with greater physical discomfort.

## Method

### Participants

With an expected large effect size, the target number of 24 participants was adapted from a similar study by Raduntz & Meffert (2019), who found significant results with the sample size of 24. Healthy adults were recruited through social media and leaflet advertising in Glasgow. Due to time constraints, the final sample fell just short of the 24 participants and included 12 participants in the experimental and 9 in the control condition. All participants’ demographic characteristics are shown in **Table 1**.

**Table 1.** Demographic Information about Participants.

*Demographic Information about Participants*
|  | Control Group |  | Experimental Group |  |
| --- | --- | --- | --- | --- |
| Variable | N (%) |  | N (%) |  |
| Gender F | 7 (78%) |  | 6 (50%) |  |
| Gender M | 1 (11%) |  | 5 (42%) |  |
| Gender NB | 1 (11%) |  | 1 (8%) |  |
| Variable | M | SD | M | SD |
| Age | 24 | 4.95 | 27.5 | 6.47 |
| Head diameter | 55.39 | 1.62 | 56.39 | 2.16 |

Means (M) and standard deviations (SD) of the demographic data of the participants. F = females; M = males, NB = non binary; N = number of participants; % = percentage of participants.

### Materials/Stimuli

#### Behavioural measures

State fatigue was evaluated using the Fatigue subscale of the Visual Analogue Scale for fatigue (VAS) consisting of 13 items, that assessed the participants’ current fatigue by asking them to mark between two extremes on a continuous line from 0 to 100 (e.g., “not at all” and “extremely”). The scale has a high reliability with Cronbach’s α ranging between .94 and .96. (Lee et al., 1991). The total change in fatigue severity score was calculated as the difference between the post-experiment mean score and initial mean score (i.e. the mean change in fatigue).

EEG headset discomfort (EEGhd) was assessed using a set of questionnaires measuring wearing comfort, pain intensity, pain distress and head muscoskeletal discomfort. The wearing comfort questionnaire was adapted from Raduntz & Meffert’s (2019) in that the two items “wearing comfort” (reverse coded) and “wearing time” were used in this study. Wearing comfort was ranked between 1 (the most appropriate) and 7 (the least appropriate). The maximal estimated tolerable wearing time of the EEG headset was obtained from a scale extending from 0 to 120 minutes with 5 minute steps.

The measurement of change in pain intensity and pain distress was based on De Pascalis et al. (2019). Pain intensity was again assessed on a Visual Analogue Scale (0 – 100) (Jensen et al., 1986). Participants were asked to rate the current perceived pain intensity (0 = no pain, 100 = maximum pain tolerable). Pain distress was measured on a 11-item rating scale (0 = neutral, 10 = worst distress imaginable).

Muscoskeletal head discomfort items were derived from the Cornell Muscoskeletal Discomfort Questionnaire (CMDQ) (Hedge et al., 1999). CMDQ addresses the intensity, frequency and task interference of muscoskeletal discomfort experienced in 20 body parts on a 3-item rating scale. For this study, the questionnaire was adjusted to address intensity and task interference of muscoskeletal discomfort in 7 head parts (crown of the head, back of the head: right, back of the head: left, neck, forehead, right ear, left ear). Internal consistency of the new items was high with Cronbach’s α = .86.

The upper and lower limits of all scales were standardized (0 – 100). The overall score capturing the EEGhd was determined as the average score of the following items: physical discomfort rating related to wearing the EEG headset; change in pain intensity and pain distress scores measured before and after the experiment; mean score from the adjusted CMDQ. The Cronbach’s α of .77 revealed satisfactory internal consistency of the four items composing the overall EEGhd score.

#### Neurofeedback device

The EEG neurofeedback device used in this study was the NeXus-10 MKII amplifier (Mind Media, BV, The Netherlands). Two experimental electrodes were located following the 10-20 placement system (Marzbani et al., 2016). Alpha activity was recorded from the occipital lobe, using gel electrodes located at O1 and O2. The ground electrode was placed over Cz. In the experimental group, reference electrodes were placed on both ears (A1 and A2), using ear clips, and a headband was used to secure the experimental electrodes. In the control group, reference electrodes were located at C3 and C4, and no headset was used. The neurofeedback software BioTrace+ (Mind Media BV) was used to view and store the recorded EEG signals.

#### Video task

Participants watched a 48 min long documentary video, Our Planet: Forests (Wilson, 2019) on a computer screen. The video was chosen as an emotionally neutral task. Nature documentary videos have been used previously in EEG research as a control task (Brusa et al., 2021).

### Procedure

Participants were seated in front of a computer screen. At the start, they completed a set of questionnaires: the consent form, demographic information, the VAS fatigue questionnaire as well as the pain and pain distress questionnaires. Head diameter in cm was measured above the participants’ eyebrows using a tape measure. Subsequently, the EEG electrodes were placed on the scalp using a conductive gel. In the experimental group, ear clips and a headband were used. Participants were instructed to watch the documentary video attentively, while their brain activity was recorded.

Following the video, participants answered three simple control questions about the video content (1: first half, 2: middle, 3: second half). Furthermore, they completed the VAS fatigue questionnaire and the EEGhd set of questionnaires while wearing the EEG headset. Finally, after removing the headset (or electrodes), participants provided qualitative feedback about their experience. The whole session lasted approximately 80 minutes.

### Statistical Analyses

A two-sample t-test was conducted in R version 4.1.2 (RStudio Team, 2021) with packages Tidyverse (Wickham et al., 2019) and lsr (Navaro, 2015). Separate t-tests examined the mean EEGhd score and the mean fatigue score change, comparing the experimental group to the controls. The assumptions for the two-sample t-test were met: the two samples were independent; there were no extreme outliers and the data was normally distributed, as confirmed per Shapiro-Wilk normality test; a Levene’s test revealed that the variance between the two groups was significantly different, therefore, the Welch’s t-test was used. Cohen’s D was used to determine the effect size.

A Pearson correlation test was carried out to examine the relationship between head diameter score and physical discomfort at the back of the head in the experimental group. For this purpose, CMDQ rating scores from “back of the head: left” and “back of the head: right” were combined to reflect a larger area at the back of the head. Additionally, a Pearson correlation between EEGhd and fatigue score change was conducted using data from all participants.

## Results

The mean scores of the behavioural variables in the experimental and the control group are summarised in **Table 2**.

**Table 2.**
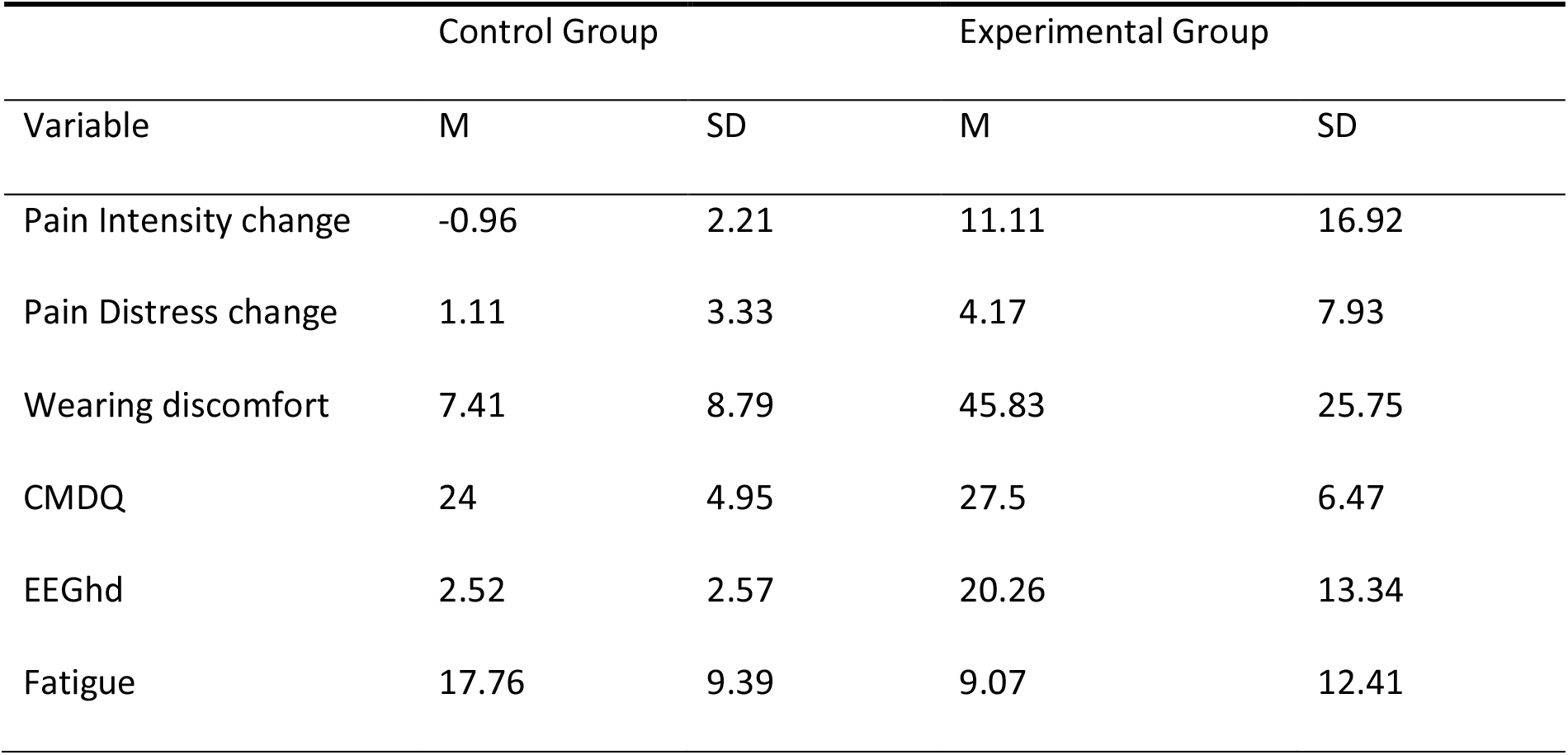
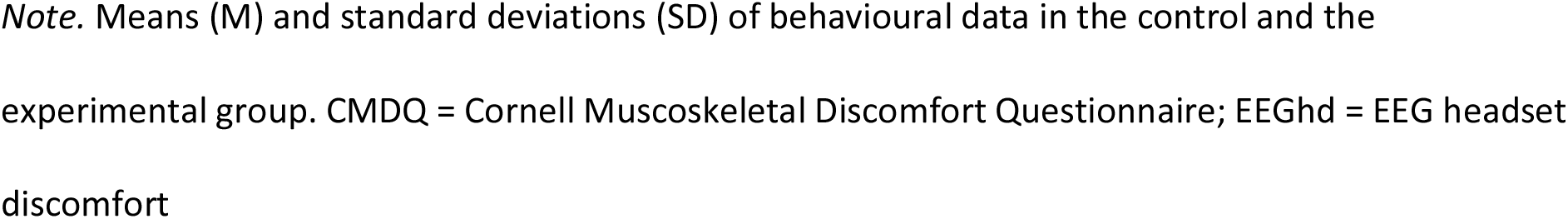
Table showing means and standard deviations of the behavioural measures.

| Variable | Control Group |  | Experimental Group |  |
| --- | --- | --- | --- | --- |
|  | M | SD | M | SD |
| Pain Intensity change | -0.96 | 2.21 | 11.11 | 16.92 |
| Pain Distress change | 1.11 | 3.33 | 4.17 | 7.93 |
| Wearing discomfort | 7.41 | 8.79 | 45.83 | 25.75 |
| CMDQ | 24 | 4.95 | 27.5 | 6.47 |
| EEGhd | 2.52 | 2.57 | 20.26 | 13.34 |
| Fatigue | 17.76 | 9.39 | 9.07 | 12.41 |

*Note.* Means (M) and standard deviations (SD) of behavioural data in the control and the experimental group. CMDQ = Cornell Musculoskeletal Discomfort Questionnaire; EEGhd = EEG headset discomfort

The mean EEGhd score of the experimental group (M = 20.26; SD = 13.34) and the control group (M =2.52; SD = 2.57) are shown in **Figure 1**. The Welch two-sample t-test showed a significant difference between the groups, *t*(12.08) = -4.5, p < .001 with a large effect size of d = 1.85 (Cohen, 1992). This result indicated higher EEGhd in the experimental than the control group confirming the hypothesis that wearing the EEG headset for a prolonged time caused significant physical discomfort.

**Figure 1.**
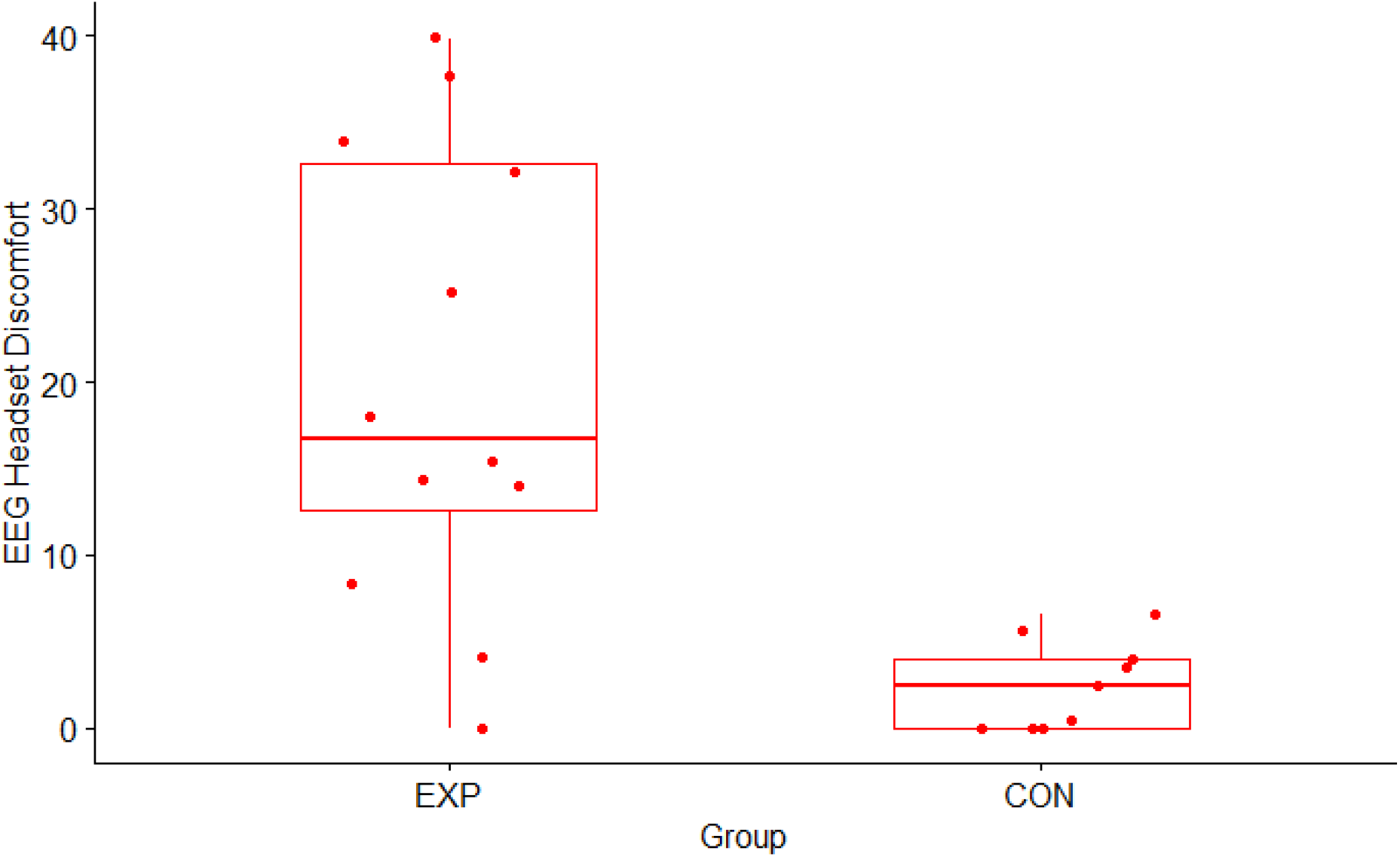
Distribution of the EEG headset discomfort across groups. *Note*. Boxplots showing the distribution of EEG headset discomfort scores across groups. EXP = experimental group; CON = controls.

There was no significant difference in the fatigue change between the groups based on the Welch’s two-sample t-test (p = .19). Furthermore, the Pearson correlation tests did not reveal any significant relationship between EEGhd and fatigue (p = .54), nor physical discomfort at the back of the head and head diameter (p = .12).

## Discussion

The main purpose of this preliminary study was to if wearing the recommended neurofeedback EEG headset for 1 hour would cause physical discomfort and mental fatigue. As predicted, our results revealed a significant difference in the EEGhd score between the experimental (wearing the headset) and the control group (not wearing the headset), showing that wearing the EEG headset for 1 hour induced significant discomfort.

This result extends previous findings that rigid EEG headsets are associated with wearing discomfort (Raduntz & Meffert, 2019). This is likely due to the pressure created by the fixed-sized headband and the ear clips (Verwulgen et al., 2018). The finding is at odds with a study that found headband EEG headsets comfortable for patients (Jochumsen et al., 2020). However, the wearing time of the EEG headset in that study was 15 minutes only. In the current study, most participants’ qualitative feedback included reports of increasing pressure and discomfort after 25 minutes (middle of the video) from the start of the experiment. Furthermore, some participants felt that they were not able to concentrate on the video due to the persistent ache. This is in line with research showing a link between physical discomfort and worsened cognitive task performance (DenHartog & Koerhius, 2017). Therefore, it is important to consider EEGhd as a factor that may interfere with the effects of EEG neurofeedback training.

Fatigue has been found previously to interfere with brain-computer interface (BCI) performance (Myrden & Chau, 2015). In the current study, the average change in fatigue severity did not differ significantly across the two groups. Also anecdotally, both groups reported an increase in fatigue due to the relaxing nature of the documentary, irrespective of the EEG headset. Hence, the current results did not show an association between EEGhd and fatigue: despite inducing discomfort, the EEG headset did not cause fatigue.

The relationship between head diameter and discomfort at the back of the head was not significant. This does not align with previous research by Nijober et al. (2015) who reported increased physical discomfort among individuals with larger head sizes. However, in the current study, subjects with a head diameter above the mean (mostly males and non-binary) were more likely to report negative experiences with the headset, using words such as “burning pain”, “persistent ache” or “discomfort”. Female participants, on average, had a smaller head diameter than males. Therefore, the headband created less pressure and discomfort. Nevertheless, the literature indicates that women are more sensitive to pain than men (Bartley & Fillingim, 2013; De Pascalis et al., 2019). This manifested itself in the present study in so far that females (but not males or non-binary) reported discomfort at both ears due to the ear clips.

The results should be interpreted with caution due to the small sample size. There was a large variance in the EEGhd scores among the experimental group, when compared to controls. This may be due to the higher variability of the experimental group both in terms of gender and age (Rusticus & Lovato, 2014). Furthermore, despite significantly higher levels of comfort in the control group, the current study did not compare the quality of the EEG alpha signal of the two groups. Therefore, it is possible that the signal quality was affected by insufficient contact with the head and non-standard placement of the reference electrodes.

The present findings provide important information about the changes in physical discomfort due to wearing an EEG headset for 1 hour. This data will be useful in the planning of an EEG neurofeedback rehabilitation study, as physical discomfort has been found to interfere with the power of the specific EEG frequencies (DePascalis et al., 2019; Shao et al., 2012), as well as the cognitive performance (DenHartog & Koerhius, 2017). Adjustable headbands are recommended for future EEG studies, as they would reduce discomfort without affecting the signal quality. Additionally, future studies should investigate alternative placements of reference electrodes to reduce the discomfort from ear clips while maintaining high quality EEG data. Furthermore, the EEGhd measure designed specifically for this study showed good internal consistency. It should be tested for validity and test-retest reliability, to increase its potential as a tool to assess physical discomfort of EEG headsets in future studies.

## Conclusion

This preliminary study is the first controlled study to investigate physical discomfort and fatigue in relation to the wearing of an EEG headset, intended for future use for neurofeedback rehabilitation of post-stroke fatigue. Results showed that wearing the EEG headset caused significantly greater discomfort than wearing electrodes without the headset, while the difference in fatigue was not significant. We also showed that the new EEG headset discomfort measure could be used to identify sources of discomfort. In the future, this would help to identify aspects of a participants’ experience that may interfere with the effects of the EEG neurofeedback rehabilitation. An adjustable headband creating less pressure rather than a rigid headset should be considered instead.

## Data Availability

All data produced in the present study are available upon request to the authors.

